# Risk of hospitalization and mortality after breakthrough SARS-CoV-2 infection by vaccine type and previous SARS-CoV-2 infection utilizing medical claims data

**DOI:** 10.1101/2021.12.08.21267483

**Authors:** Meghana Kshirsagar, Sumit Mukherjee, Md Nasir, Nicholas Becker, Juan Lavista Ferres, Barbra A. Richardson

**Affiliations:** AI for Good Research Lab, Microsoft Corporation, Redmond, WA 98052, USA; Departments of Biostatistics and Global Health, Univ. of Washington, Seattle, WA 98195, USA; Vaccine and Infectious Disease Division, Fred Hutch Cancer Research Center, Seattle, WA 98109, USA

**Keywords:** breakthroughs, vaccines, Pfizer, Moderna, Janssen, SARS-CoV-2, COVID-19

## Abstract

We compare the risks of hospitalization (n=1121) and mortality (n=138) in a cohort of 17,881 breakthrough SARS-CoV-2 infections for the Pfizer, Moderna and Janssen vaccines for those with and without SARS-CoV-2 infections prior to vaccination. Cox regression analysis results in a lower hazard ratio for breakthroughs receiving the Moderna vaccine, but a significantly higher hazard ratio for breakthroughs receiving the Janssen vaccine, as compared to breakthroughs who got the Pfizer vaccine. Further, the risk of hospitalization (P<0.001) and death (P<0.05) were lower among breakthroughs who had a SARS-CoV-2 infection prior to vaccination, independent of age, sex, comorbidities, and vaccine type.

**Note:** We do not study the role of natural immunity in SARS-CoV-2 infections, as our cohort does not contain unvaccinated individuals.

## Background

The combination of widespread COVID-19 vaccination and high community levels of SARS-CoV-2 circulating throughout the United States has led to many breakthrough SARS-CoV-2 infections [1,4,5]. Breakthrough infections are generally less severe than infections in unvaccinated individuals [6,7]. While the risk of breakthrough SARS-CoV-2 infection and COVID-19 mortality have recently been reported by type of vaccine [8], little to no information exists regarding risk of hospitalization by vaccine type for breakthrough infections [1]. In addition, while prior SARS-CoV-2 infection is associated with a lower risk of breakthrough infection, it is unknown how large an effect a prior infection has on the severity of breakthrough COVID-19 infections [3].

COVID-19 vaccinations began in the United States in late December 2020. By late February 2021, the Pfizer-BioNTech (Pfizer), Moderna, and Johnson & Johnson / J&J (Janssen) vaccines were all approved for emergency use authorization (EUA). We used de-identified United States medical claims records from Change Healthcare to estimate the risk of hospitalization and of death, by vaccine type and by previous SARS-CoV2 infection, among SARS-CoV-2 breakthrough infections in those individuals fully vaccinated between Mar 10^th^, 2021 and April 27^th^, 2021.

## Methods

### Ethics approval and consent to participate

This study does not constitute as human subjects research due to the usage and reporting of only deidentified observational data as determined by the ethics committee of the University of Washington School of Medicine. An ethics approval waiver and a consent waiver were received from the ethics committee of the University of Washington School of Medicine.

### Study population

Our study uses claims records from Change Healthcare collected over a period from March 1, 2020, to Sept 30, 2021, encompassing over 100 million records from over 8 million patients. This dataset includes all COVID-19 positive patients, identified by the ICD-10 diagnosis codes of U07.1 (COVID-19, virus identified, lab confirmed) as the principal diagnosis. From this cohort of 8.18 million patients, fully vaccinated individuals are identified by looking for procedure codes encoding the second doses of Pfizer (0002A) and Moderna (0012A), and the first dose of Janssen (0031A). Amongst these individuals, breakthrough patients were defined as those who had a COVID-19 diagnosis at least 14 days after the date of vaccination (See Supplementary Figure 1 for the criteria used for cohort selection). The Pfizer and Moderna vaccination drives started much earlier, in late December (Supplementary Figures 2 and 3), as compared to that of the Janssen vaccines, which also saw a stall in vaccine rates in mid-April (Supplementary Figure 4). Based on these observations, our cohort consisted of individuals who were fully vaccinated between Mar 10^th^, 2021 – Apr 27^th^, 2021, the period during which all three vaccines were being widely administered. These were followed from the date of vaccination up to Oct 14^th^, 2021.

### Statistical methods

Date of full vaccination was defined as 14 days after: 1) a single Janssen vaccine; 2) the second Moderna vaccine dose; or 3) the second Pfizer vaccine dose. Cox proportional hazards regression was used to estimate univariate hazard ratios (HRs) and multivariable HRs in a model including age (categorized), sex, vaccine type, Elixhauser comorbidities (encoded as independent binary variables), and previous SARS-CoV-2 infection. Interactions between vaccine type and all other covariates and previous infection and all other covariates were tested but none were statistically significant. All analyses were performed using the ‘coxph’ function from the R package ‘survival’ [11].

## Results

Our study includes 17,881 fully vaccinated patients with breakthrough SARS-CoV-2 infections between March 10^th^ and Oct 14^th^, 2021. Of those patients, 10,011 received Pfizer, 5,028 received Moderna, and 2,842 received Janssen. Breakthrough cases receiving Janssen were younger than those receiving Pfizer or Moderna and were slightly more likely to be male (Table 1). Breakthrough cases receiving Moderna were more likely to have had COVID-19 prior to vaccination.

**Table 1.** Characteristics of SARS-CoV-2 Breakthrough Infections Cohort tracked from March 10, 2021 to Oct 14^th^, 2021

| Variable | Pfizer<br>n=10,011 | Moderna<br>n=5,028 | Janssen<br>n=2,842 | Overall<br>n=17,881 |
| --- | --- | --- | --- | --- |
| Age |  |  |  |  |
| 0-20 | 98 (1.0%) | 37 (0.7%) | 42 (1.5%) | 177 (1.0%) |
| 20-35 | 947 (9.4%) | 462 (9.2%) | 349 (12.3%) | 1,758 (9.8%) |
| 35-50 | 1,647 (16.4%) | 777 (15.5%) | 712 (25.1%) | 3,136 (17.5%) |
| 50-64 | 3,269 (32.6%) | 1,483 (29.5%) | 1,148 (40.4%) | 5,900 (33.0%) |
| 64-80 | 3,391 (33.9%) | 1,826 (36.3%) | 500 (17.6%) | 5,717 (32.0%) |
| >80 | 659 (6.6%) | 443 (8.8%) | 91 (3.2%) | 1,193 (6.7%) |
| Male | 4,462 (44.5%) | 2,182 (43.4%) | 1,322 (46.5%) | 7,966 (44.6%) |
| Prior SARS-CoV-2<br>Infection | 1,222 (12.2%) | 1,078 (21.4%) | 349 (12.3%) | 2,649 (14.8%) |

Table 1.
| Variable | Hospitali-<br>zation<br>Incidence<br>Per 100<br>Person<br>Years<br>(n/pys) | Hospitalization<br>Univariate HR<br>(95% CI)<br>n=17,881 | Hospitalization<br>Multivariate aHR<br>(95% CI)<br>n=17,881 | Mortality<br>Incidence<br>Per 100<br>Person<br>Years<br>(n/pys) | Mortality<br>Univariate HR<br>(95% CI)<br>n=17,881 | Mortality<br>Multivariate aHR<br>(95% CI)<br>n=17,881 |
| --- | --- | --- | --- | --- | --- | --- |
| Vaccine |  |  |  |  |  |  |
| <u>Pfizer</u> | 20.1 | 1.0 | 1.0 | 2.6 | 1.0 | 1.0 |
| Moderna | 19.2 | 0.93 (0.8--1.1) | 0.85 (0.74--0.99)* | 2.3 | 0.89 (0.61--1.3) | 0.98 (0.6--1.5) |
| Janssen | 26.5 | 1.69 (1.5--2.0)*** | 1.78 (1.51--2.09)*** | 3.0 | 1.47 (0.98--2.2) | 1.70 (1.0--2.8)* |
| Age |  |  |  |  |  |  |
| 0-20 | 1.9 | 0.29 (0.04--2.1) | 0.34 (0.05--2.43) | 1.9 | 7.48 (0.8--71.9) | 8.59 (0.9--83.4) |
| 20-35 | 1.9 | 0.27 (0.15--0.5)*** | 0.30 (0.16--0.56)*** | 0.0 | 0.0 (–) | 0.0 (–) |
| <u>35-50</u> | 6.8 | 1.0 | 1.0 | 0.3 | 1.0 | 1.0 |
| 50-64 | 16.9 | 2.08 (1.6--2.7)*** | 1.94 (1.51--2.49)*** | 1.8 | 5.82 (1.8--18.9)** | 4.77 (1.4--15.8)* |
| 64-80 | 31.7 | 2.96 (2.3--3.7)*** | 2.81 (2.20--3.59)*** | 3.9 | 12.30 (3.9-38.9)*** | 9.06 (2.8-29.4)*** |
| >80 | 52.9 | 4.35 (3.3--5.7)*** | 3.93 (2.97--5.20)*** | 9.1 | 24.60 (7.6-79.7)*** | 17.10 (5.0-58.2)*** |
| Sex |  |  |  |  |  |  |
| <u>Female</u> | 17.5 | 1.0 | 1.0 | 2.2 | 1.0 | 1.0 |
| Male | 25.0 | 1.38 (1.2--1.5)*** | 1.23 (1.09--1.39)*** | 3.0 | 1.26 (0.9-1.7) | 1.10 (0.8--1.6) |
| Prior<br>SARS-CoV2<br>Infection |  |  |  |  |  |  |
| <u>No</u> | 21.9 | 1.0 | 1.0 | 2.7 | 1.0 | 1.0 |
| Yes | 7.5 | 0.56 (0.4--0.8)*** | 0.49 (0.35--0.69)*** | 0.5 | 0.21 (0.05-0.84)* | 0.24 (0.06--0.98)* |

\*P<0.05
\*\*P<0.01
\*\*\*P<0.001

Risk of hospitalization and mortality among breakthrough cases increased with older age and was higher for male patients (Table 2). In multivariable analyses controlling for age, male sex, comorbidities, and prior SARS-CoV-2 infection, the risk of hospitalization was lower for breakthrough cases receiving the Moderna vaccine (adjusted Hazard Ratio (aHR): 0.85, 95% Confidence Interval (CI) 0.74--0.99, p<0.05) but was significantly higher for breakthrough cases who received the Janssen vaccine compared to breakthrough cases who received the Pfizer vaccine (aHR: 1.78, 95% CI 1.51--2.09, p<0.001). There was an increase in the rate of hospitalization amongst breakthrough cases starting ∼110-125 days after full vaccination for all three vaccines depending on age group, with a steeper increase for Janssen (Supplementary Figure 5 and 6). The comorbidities with statistically significant hazard ratios for breakthrough SARS-CoV-2 infection include lung disease, cancers, hypertension, coagulopathy, renal failures, alcohol abuse, anemia, seizures, and arthritis (Supplementary Table 1).

Risk of mortality for breakthrough cases receiving Pfizer and Moderna vaccines was similar, but higher for Janssen recipients compared to Pfizer (aHR: 1.70, 95% CI 1.03–2.80, p<0.05). Finally, breakthrough cases who had a previous SARS-CoV-2 infection were half as likely to be hospitalized (aHR: 0.49, 95% CI 0.35—0.69, p<0.001) and four times less likely to die (aHR: 0.24, 95% CI 0.06--0.98, p<0.05), when compared to breakthrough cases without a prior SARS-CoV-2 infection independent of age, sex, comorbidities, and vaccine type.

The time to hospitalization in breakthrough cases shows a steady increase over the 7 months assessed (Supplementary Figure 7), suggesting that vaccines were effective over long periods of time. While the number of hospitalizations increased exponentially in the Delta predominance period between June 2021 and Aug 2021 (Supplementary Figure 8), the upward trend in the time to hospitalization was not affected during this period and largely depended on when most vaccinations took place (Mar – May).

## Discussion

Using medical claims data, we found that the risk of hospitalization in SARS-CoV-2 breakthrough infections was lower for those receiving the Moderna vaccines, but significantly higher in breakthrough cases receiving the Janssen vaccine, compared to breakthrough cases who received the Pfizer vaccine. The risk of mortality was similar in breakthrough infections who received the Pfizer and Moderna vaccines, but higher in breakthrough infections receiving Janssen vaccine, compared to breakthroughs receiving the Pfizer vaccine. These findings are similar to those reported by the CDC for mortality but provide additional information regarding risk of hospitalization by vaccine type [1,2]. We also found older age, male sex, and certain comorbidities to be risk factors for more severe breakthrough infections, which is similar to what has been reported in prior studies of SARS-CoV-2 infections among unvaccinated individuals [9].

Importantly, we found that risk of hospitalization was 50% less, and risk of death was 75% less, among breakthrough cases who had a SARS-CoV-2 infection prior to vaccination than that for breakthrough cases without a previous infection. While other studies have reported lower risk of breakthrough infection with previous SARS-CoV-2 infection, our study shows that immunity provided by previous infection seems to provide additional protection, over that provided by vaccines, against severe COVID-19 independent of vaccine type, age, comorbidities, and sex [3].

An important strength of our study is that we consider US-wide breakthrough hospitalizations covering a broad demographic, and compare all three vaccines, whereas most previous studies lack specific data on Janssen. Limitations of our study include, first, a lack of access to data on unvaccinated individuals or those that had a negative SARS-CoV-2 test result. Second, the medical claims data that our cohort comes from, consists of mostly privately insured individuals and is thus likely to miss people with the most adverse outcomes.

Our findings add to the growing literature regarding SARS-CoV-2 breakthrough infections and protection provided by previous SARS-CoV-2 infections against severe disease. This study reinforces the need for booster vaccination shots to protect against more severe COVID-19 among those initially receiving the Janssen vaccine and provides new information regarding the role of prior SARS-CoV-2 infection and lower risk of more severe breakthrough infections.

## Data Availability

All data produced in the present study are available upon reasonable request to the authors

## Notes

The authors do not have an association that might pose a conflict of interest

### Competing Interest Statement

The authors have declared no competing interest.

### Funding Statement

This study did not receive any funding

### Author Declarations

Ethics approval and consent to participate This study does not constitute as human subjects research due to the usage and reporting of only deidentified observational data as determined by the ethics committee of the University of Washington School of Medicine. An ethics approval waiver and a consent waiver were received from the ethics committee of the University of Washington School of Medicine.

### Summary of Updates

Updated the wording in the abstract, intro and conclusion to clear the confusion surrounding the prior-covid results

