## Supplementary information for "Risk of hospitalization and mortality after breakthrough SARS-CoV-2 infection by vaccine type and previous SARS-CoV-2 infection utilizing medical claims data" (from another version of this preprint: version 1, 2021-12-09)

### **Contents**

Figure 1: Study cohort and vaccine-level statistics of the population

Table 1: Adjusted hazards ratio (aHR) of the 39 Elixhauser comorbidities, for Hospitalization and Mortality After Breakthrough SARS-CoV-2 Infection, estimated from Cox proportional hazards models.

Figure 2: Distribution of vaccination dates in individuals getting Pfizer vaccine’s second dose.

Figure 3: Distribution of vaccination dates in individuals getting Moderna vaccine’s second dose.

Figure 4: Distribution of vaccination dates in individuals getting Janssen vaccine.

Figure 5: Cox survival plots showing rate of hospitalization for the age group: (35,50] for all three vaccines.

Figure 6: Cox survival plots showing hospitalization-free survival for the age group: (50, 64] for all three vaccines. We show the curves for the crosshatch between gender and prior COVID-exposure.

Figure 7: Average time to hospitalization shown for every month in the breakthrough cases arising in individuals who took the Pfizer or Moderna vaccines. We use all vaccination data from Jan 5^th^, 2021 to Oct 14^th^, 2021 and restrict this analysis to Pfizer and Moderna where we have reliable data over this period.

Figure 8: The number of people hospitalized with breakthrough SARS-CoV-2 infections by month and vaccine type

Figure 1: Study cohort and vaccine-level statistics of the population

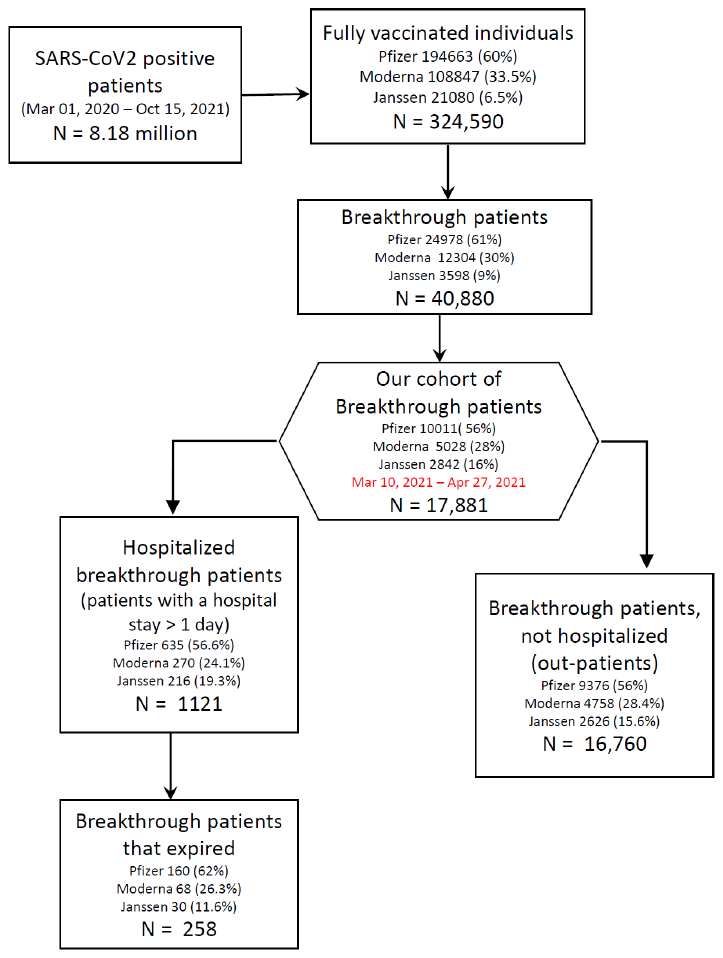

Table 1: Adjusted hazards ratio (aHR) of the 39 Elixhauser comorbidities, for Hospitalization and Mortality After Breakthrough SARS-CoV-2 Infection, estimated from Cox proportional hazards models.

| Elixhauser Comorbidity | Incidence percentage (%) n/population | Hospitalization Multivariate aHR (95% CI) N=17,881 | Mortality Multivariate aHR (95% CI) N=17,881 |
| --- | --- | --- | --- |
| AIDS | 0.79 | 1.20 (0.62--2.34) | 1.35 (0.18--9.94) |
| Alcohol abuse | 2.23 | 1.61 (1.14--2.27)** | 1.90 (0.76--4.74) |
| Deficiency anaemia | 15.83 | 1.25 (1.05--1.49)* | 1.68 (1.04--2.71)* |
| Rheumatoid arthritis/collagen vascular diseases | 6.43 | 1.26 (1.02--1.55)* | 1.14 (0.62--2.09) |
| Blood loss anaemia | 1.82 | 0.85 (0.60--1.21) | 0.78 (0.30--2.05) |
| CHF (congestive heart failure) | 7.00 | 1.18 (0.97--1.43) | 1.56 (0.94--2.58). |
| Chronic Lung disease | 16.75 | 1.22 (1.05--1.42)** | 0.84 (0.54--1.30) |
| Coagulopathy | 4.37 | 1.35 (1.09--1.66)** | 1.08 (0.63--1.86) |
| Cerebrovascular disease (on admission) | 6.26 | 0.90 (0.73--1.10) | 0.73 (0.42--1.26) |
| Cerebrovascular disease SQLA | 1.20 | 1.04 (0.63--1.71) | 0.99 (0.31--3.14) |
| Depression | 14.60 | 1.06 (0.89--1.26) | 0.81 (0.49--1.35) |
| Diabetes Complicated | 14.91 | 1.11 (0.91--1.36) | 0.98 (0.57--1.70) |
| Diabetes Uncomplicated | 19.11 | 1.17 (0.97--1.41). | 1.16 (0.70--1.93) |
| DRUG_ABUSE | 2.48 | 1.00 (0.69--1.45) | 0.30 (0.04--2.20) |
| Hypertension Complicated | 11.35 | 1.22 (0.99--1.51). | 1.43 (0.83--2.46) |
| Hypertension Uncomplicated | 37.98 | 0.75 (0.64--0.88)*** | 0.59 (0.38--0.93)* |
| Hypothyroidism | 11.82 | 1.04 (0.87--1.23) | 1.03 (0.65--1.64) |
| Cancer - Leukemia | 1.08 | 1.71 (1.18--2.45)** | 3.16 (1.41--7.07)** |
| Liver disease (mild) | 8.34 | 0.92 (0.75--1.12) | 0.50 (0.26--0.98)* |
| Liver disease (severe) | 0.74 | 1.86 (1.25--2.79)** | 7.23 (2.99--17.46)*** |
| Cancer -Lymphoma | 1.03 | 1.47 (1.05--2.07)* | 1.16 (0.45--2.99) |
| Metastatic cancer | 1.17 | 1.53 (1.06--2.22)* | 2.10 (0.82--5.35) |
| DEMENTIA | 1.61 | 0.95 (0.67--1.34) | 1.07 (0.44--2.59) |
| Neurological disorders (Movement) | 2.20 | 1.12 (0.83--1.51) | 0.85 (0.35--2.06) |
| Other Neurological disorders | 3.34 | 0.98 (0.74--1.28) | 0.90 (0.44--1.83) |
| Seizures and epilepsy | 2.36 | 1.36 (1.01--1.84)* | 0.77 (0.29--2.06) |
| OBESE | 24.18 | 1.01 (0.87--1.17) | 1.05 (0.69--1.60) |
| THYROID_OTH | 5.18 | 0.81 (0.61--1.07) | 0.78 (0.35--1.76) |
| PARALYSIS | 1.40 | 0.89 (0.54--1.45) | 2.01 (0.68--5.99) |
| Peripheral vascular disorders | 10.08 | 1.01 (0.85--1.20) | 1.46 (0.94--2.27). |
| PSYCHOSES | 4.81 | 0.97 (0.72--1.29) | 0.40 (0.12--1.32) |
| Pulmonary circulation disorders | 2.17 | 1.08 (0.82--1.42) | 0.84 (0.41--1.75) |
| Renal failure (moderate) | 7.83 | 1.31 (1.08--1.59)** | 1.05 (0.64--1.72) |
| Renal failure (severe) | 4.97 | 1.69 (1.36--2.11)*** | 2.11 (1.23--3.60)** |
| CANCER (solid tumor) | 6.07 | 0.90 (0.73--1.11) | 0.71 (0.40--1.27) |
| CANCER (in situ) | 1.31 | 1.15 (0.77--1.70) | 3.52 (1.74--7.12)*** |
| Peptic ulcer disease | 1.48 | 1.06 (0.72--1.54) | 0.55 (0.16--1.91) |
| Valvular disease | 7.97 | 1.03 (0.86--1.23) | 1.34 (0.85--2.12) |
| Weight loss | 3.76 | 1.16 (0.92--1.45) | 1.71 (1.00--2.92)* |

*P<0.05

**P<0.01

***P<0.001

**Methods:**

**Data Source:**

Roughly 95% of the claims used for this study are commercial and 5% are Medicare Advantage/other types of plans. Every medical claims record contains information about the diagnoses (in the form of ICD-10 codes), the procedures performed and prescribed drugs. The claims in our dataset include primarily open claims, and a subset of closed payer claims which are normalized for analytics purposes providing sound directional insight for this study. The open claims are derived from broad based healthcare sources and consists of all the medical claims that Change Healthcare processes and for which they have the rights to use. The closed claims are derived from the payer and capture nearly all events that occur during the patient’s enrollment period.

Figure 1: Distribution of vaccination dates in individuals getting Pfizer vaccine’s second dose. At the peak of Pfizer’s vaccination drive, our data has more than 3000 people were getting vaccinated per day.

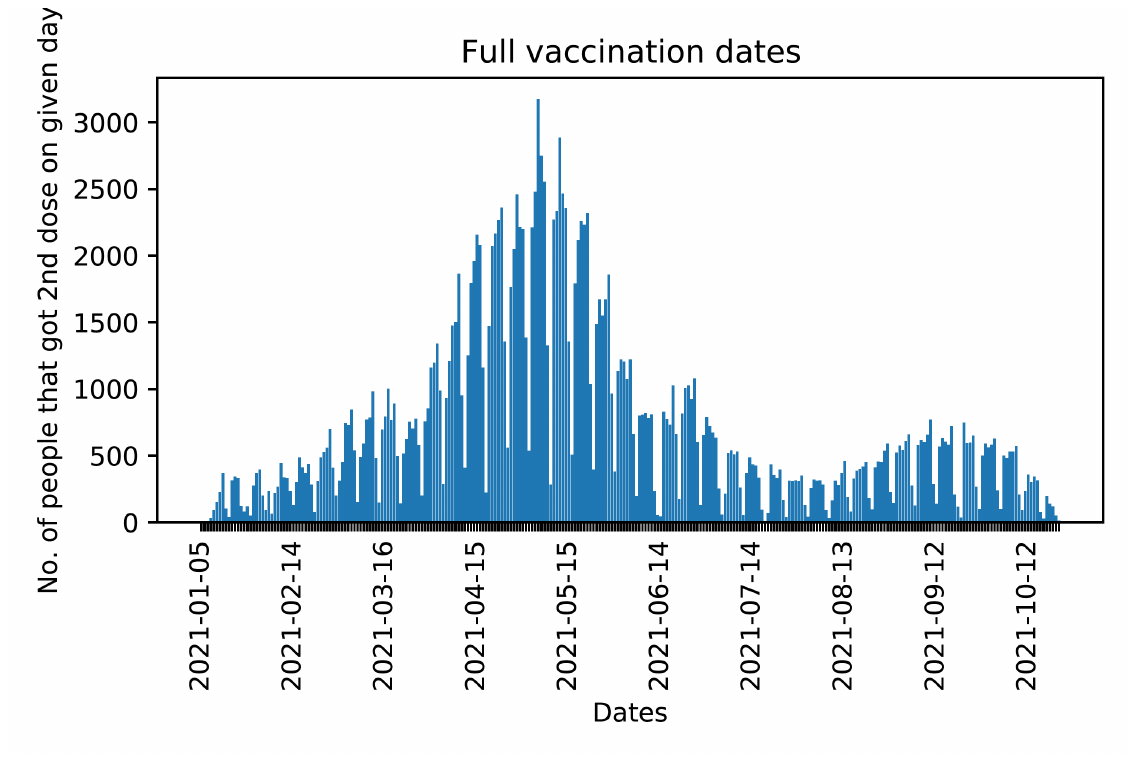

Figure 2: Distribution of vaccination dates in individuals getting Moderna vaccine’s second dose. While the distribution looks similar to that of Pfizer, the height of the peak is much lower, at around 1750.

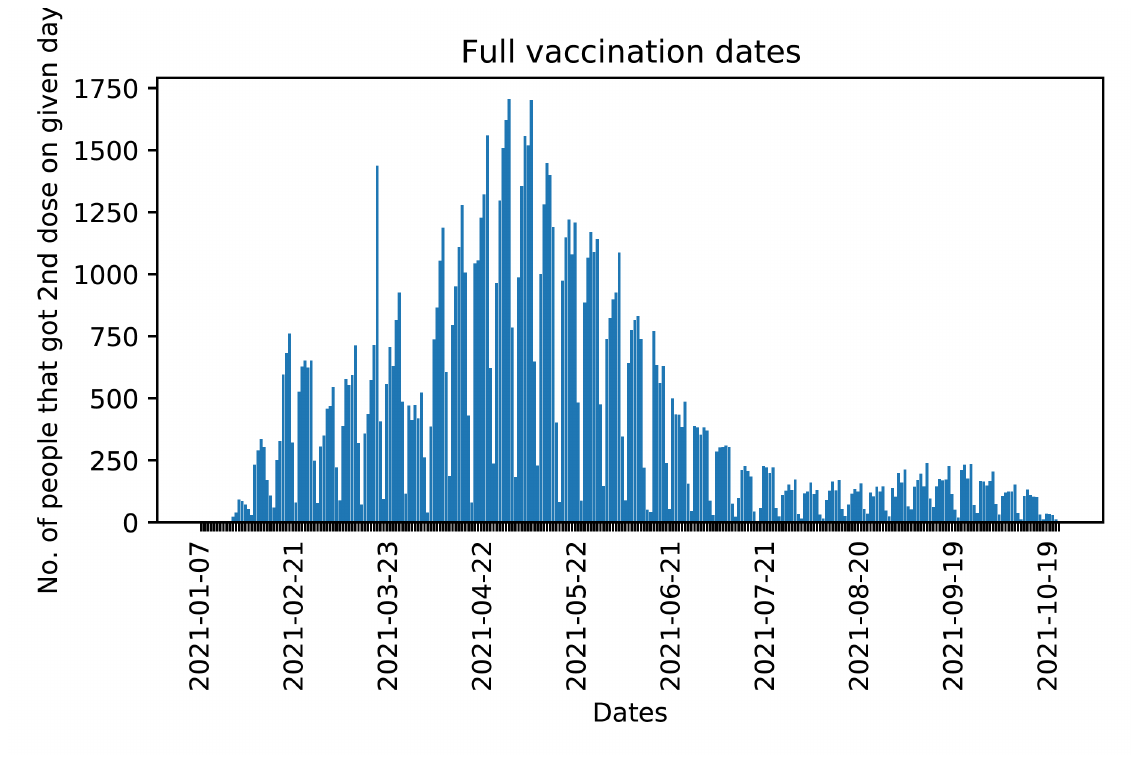

Figure 3: Distribution of vaccination dates in individuals getting Janssen vaccine, the earliest date being 24^th^ Feb, 2021. The peak is much lower at around 1000 individuals per day.

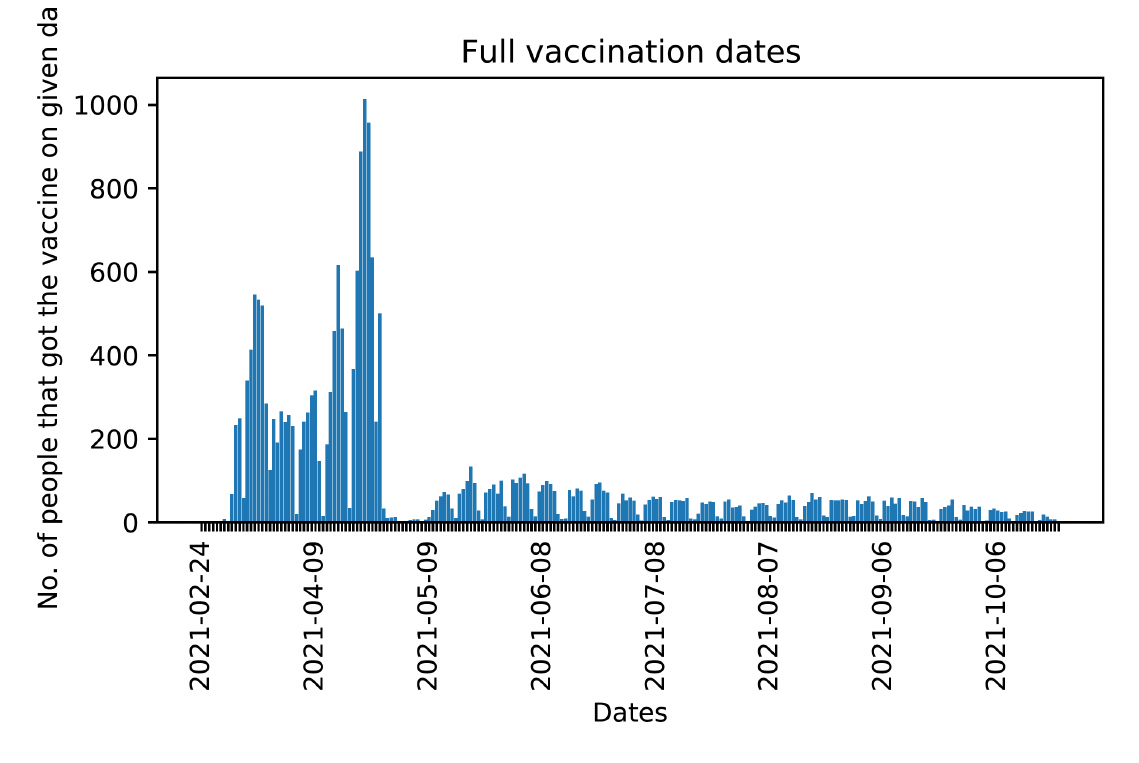

Figure 4: Cox survival plots showing rate of hospitalization for the age group: (35,50] for all three vaccines. We show the curves for the crosshatch between gender and prior COVID exposure. The lowest increase in the rate is observed for women with prior COVID exposure followed by men with prior COVID exposure starting at around 125 days, as compared to around 110 days for the other categories.

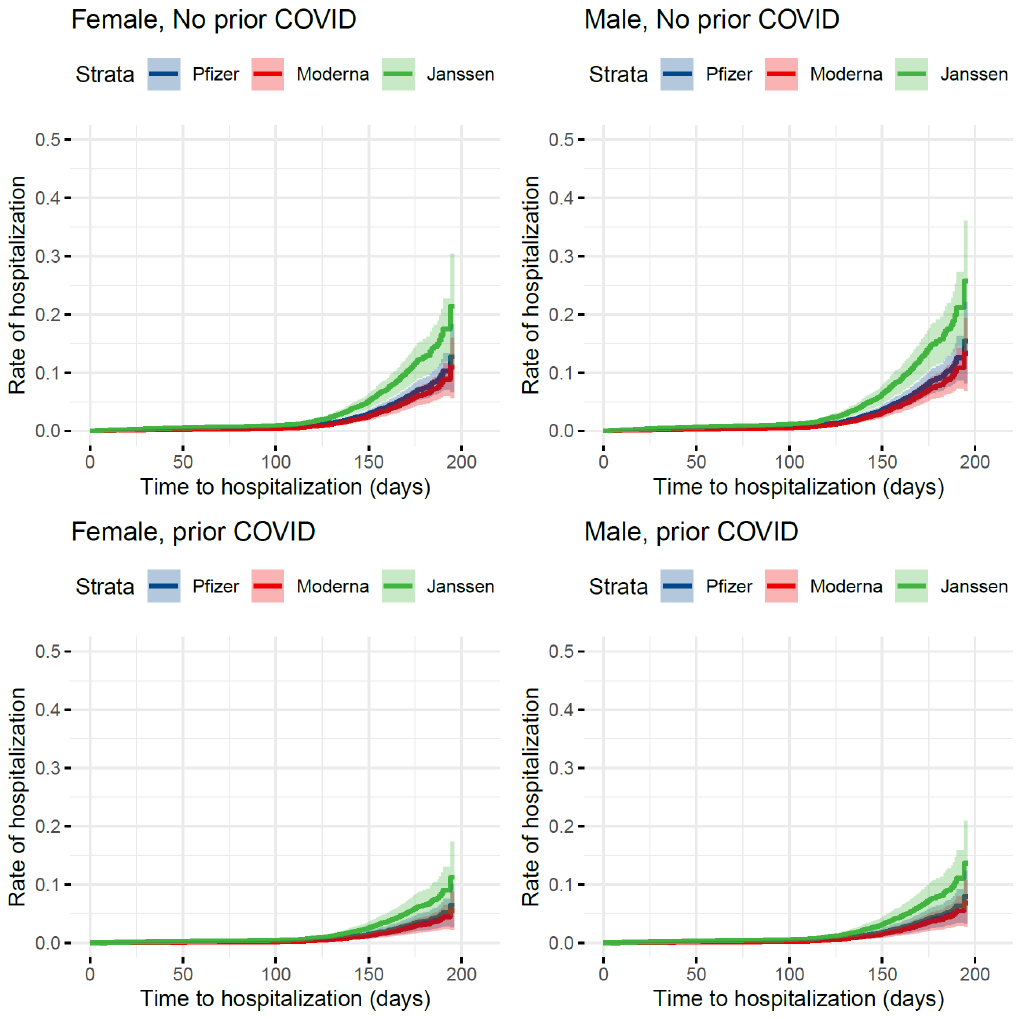

Figure 5: Cox survival plots showing hospitalization-free survival for the age group: (50, 64] for all three vaccines. We show the curves for the crosshatch between gender and prior COVID-exposure. The lowest increase in rate is observed for women with prior exposure to COVID-19, followed by men with prior COVID exposure starting at around 115 days as compared to around 100 days for men and women without prior exposure to COVID-19.

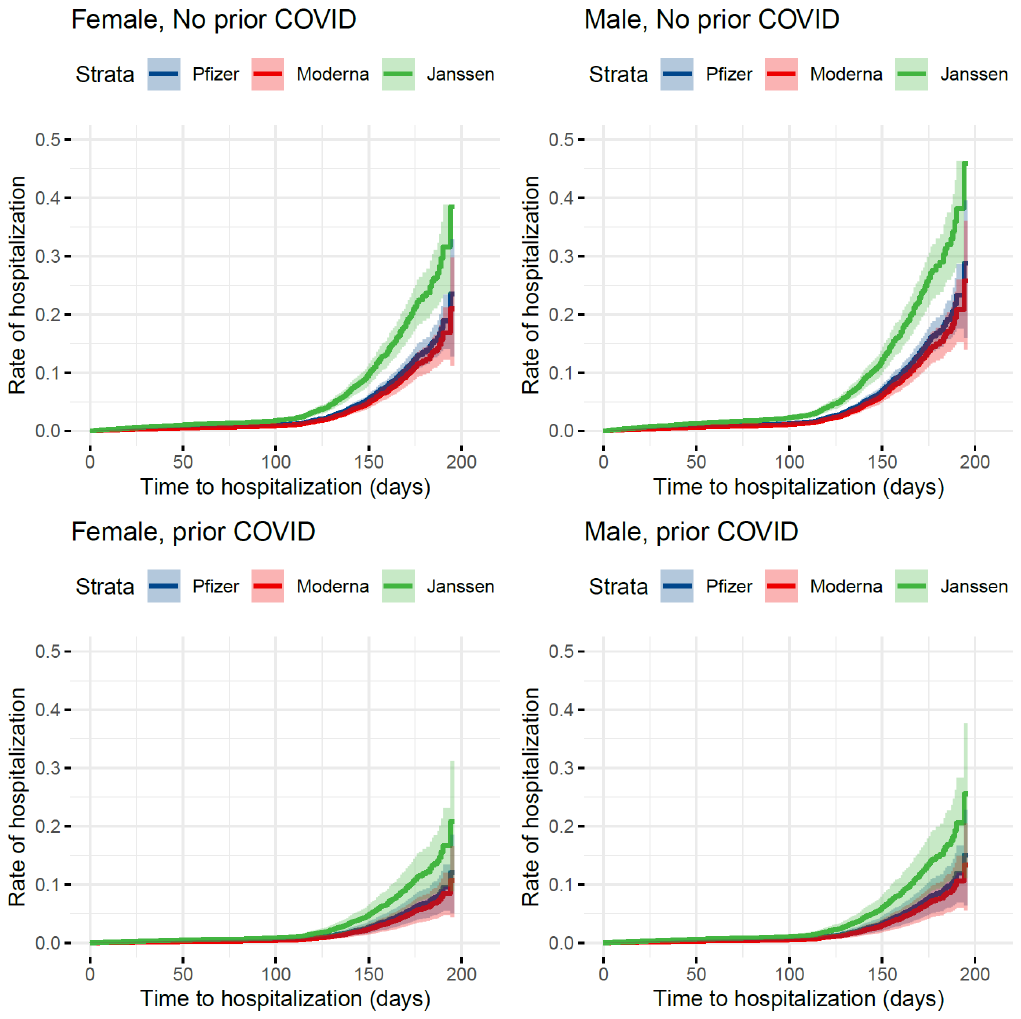

Figure 6: Average time to hospitalization shown for every month in the breakthrough cases arising in individuals who took the Pfizer or Moderna vaccines. We use all vaccination data from Jan 5^th^, 2021 to Oct 14^th^, 2021 and restrict this analysis to Pfizer and Moderna where we have reliable data over this period.

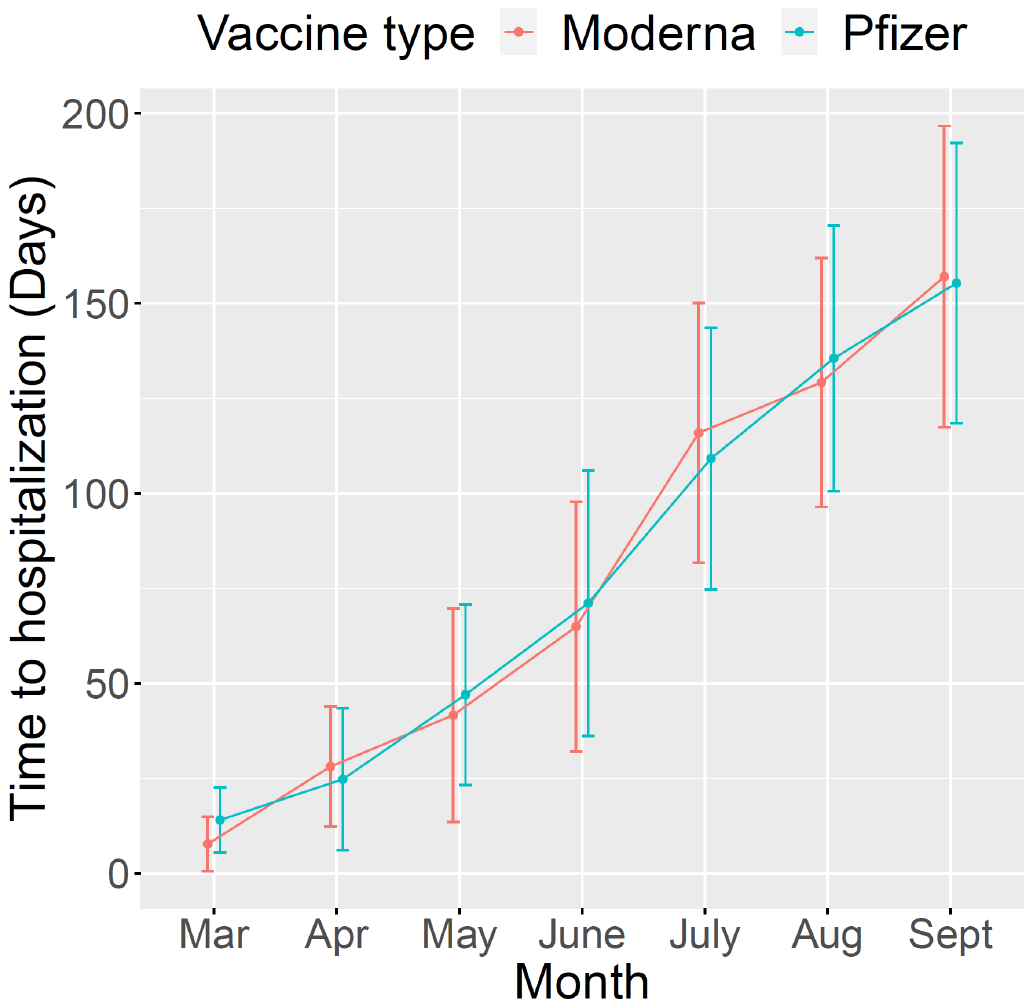

Figure 7: The number of people hospitalized with breakthrough SARS-CoV-2 infections by month and vaccine type

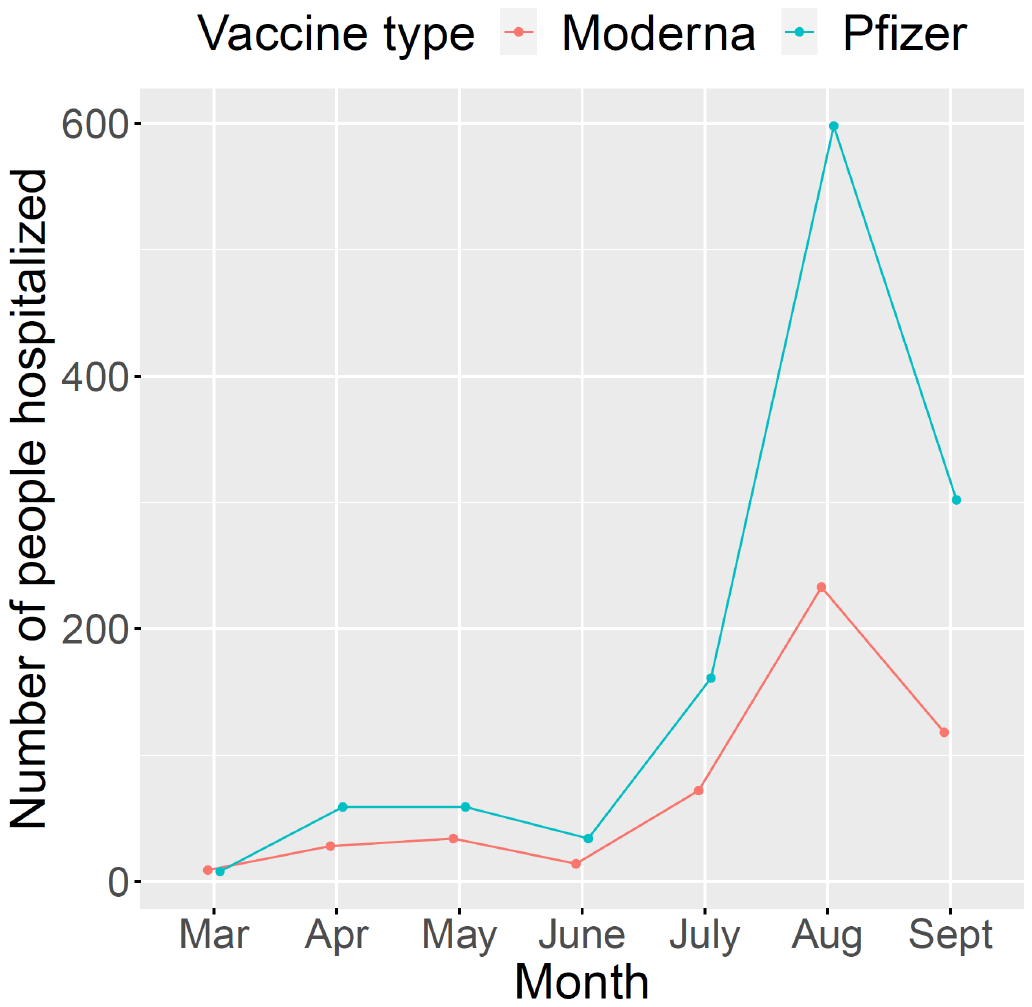
